# The interaction between chronic hepatitis B (CHB) and Metabolic dysfunction-associated steatotic liver disease (MASLD) in a diverse central London population

**DOI:** 10.64898/2026.06.15.26355674

**Authors:** Emily Martyn, Claire Mullender, Stephen Ogunnaike, Agnieszka Kemper, Indrajit Ghosh, Dimitra Peppa, Emmanoulis Tsochatzis, Richard Gilson, Stuart Flanagan, Andrew Copas, Douglas Macdonald, Alejandro Arenas-Pinto, Philippa C Matthews

## Abstract

**Introduction:** The overlap between chronic hepatitis B (CHB) and metabolic dysfunction-associated steatotic liver disease (MASLD) is an emerging global health challenge. We investigated the impact of MASLD and metabolic comorbidity in a diverse London viral hepatitis clinic.

**Methods:** This retrospective cross-sectional study (May 2018–Feb 2024) included adults with CHB having controlled attenuation parameter (CAP) measurements. MASLD was defined as CAP >264 dB/m plus ≥1 cardiometabolic factor (CMF). We used univariable and multivariable models to examine MASLD’s relationship with liver stiffness and hepatitis B viral load (HBV VL).

**Results:** Among 323 individuals (67% male, median age 36), most were from Black (35%) or non-white British/Irish (29%) backgrounds. Overall, 64% had ≥1 CMF, and 20% had MASLD. The CHB/MASLD group was significantly older (median 43 vs 35 years, p<0.001) with higher median alanine transaminase (35 vs 30 IU/L, p=0.02) and liver stiffness (5.3 vs 4.7 kPa, p<0.001). Following adjustment for covariates, MASLD remained significantly associated with liver stiffness (β = 0.48 kPa, p=0.03). While univariable analysis showed significantly lower HBV VL in people with MASLD (median 54 vs 417 IU/ml, p=0.004), adjusted multivariable analysis revealed no significant association between MASLD and log10 HBV VL (p=0.2).

**Conclusions:** Although adjusted analysis does not support an independent association between MASLD and HBV VL, the data highlight a substantial cardiometabolic burden in this CHB population and clearly link MASLD to more severe liver disease. Holistic consideration of metabolic comorbidities is crucial in comprehensive CHB management

## Introduction

Chronic hepatitis B (CHB) and metabolic dysfunction-associated steatotic liver disease (MASLD) are major causes of global liver disease. Despite an effective vaccine and suppressive antiviral treatment, CHB is responsible for almost 1 million deaths per year due to end-stage liver disease and hepatocellular carcinoma (HCC)^1^. Approximately 30% of the global population is living with MASLD, and this is projected to increase to 60% by 2050^2^. The highest MASLD prevalence is observed in Latin America, North Africa and Eastern Mediterranean - regions which also have intermediate endemicity for hepatitis B, demonstrating potential for significant disease overlap^3–5^.

Metabolic dysfunction-associated steatotic liver disease (MASLD) is defined as steatotic liver disease (SLD, >5% of the hepatocytes containing fat) occurring together with one or more cardiometabolic factor(s) (CMF(s), pre-diabetes or type 2 diabetes, dyslipidemia, hypertension, overweight or obesity) and superseded the previous term ‘non-alcoholic fatty liver disease’ (NAFLD) in 2023^6^. Existing evidence for the interaction between CHB and MASLD is conflicting, with some studies reporting an increased risk of cirrhosis and HCC in people living with CHB and MASLD, whereas other studies suggest a protective effect of MASLD^7–9^. Reasons for the inconsistency may include factors such as HBV genotype, host genetics, local environment (e.g. diet) and methodological differences between studies.

The current landscape represents a critical moment to investigate the overlap between CHB and MASLD. The World Health Organization has set ambitious targets to eliminate viral hepatitis as a public health threat by 2030, aiming for a 90% reduction in incidence and a 65% reduction in mortality^10^. Achieving these goals will require a more comprehensive understanding of factors influencing liver disease progression in people living with CHB, for example metabolic comorbidity. Furthermore, the therapeutic landscape for MASLD is rapidly evolving: resmetirom, the first pharmacological agent approved for MASLD with fibrosis, has been licensed in the United States and Europe within the past two years, while incretin-based anti-obesity therapies such as semaglutide and tirzepatide are becoming increasingly accessible^11^. Together, these developments underscore the urgency and opportunity to better define the impact of MASLD in CHB.

Our real-world, cross-sectional study aims to (a) describe and characterise cardiometabolic comorbidity in people living with CHB attending a central London viral hepatitis clinic, (b) investigate the impact of MASLD on hepatitis B outcomes using routine clinical data (e.g. viral load and laboratory/imaging assessment of liver disease). These data will contribute to the longer term goal of better tailoring personalized treatment with use of novel agents, and improve long-term outcomes.

## Methods

### Study setting and design

This is a cross-sectional retrospective real-world study based on data routinely collected at Mortimer Market Centre viral hepatitis outpatient clinic, Central and North West London NHS Foundation Trust (CNWL). The viral hepatitis service sees approximately 600 people living with CHB per year (100 of whom have HIV coinfection) and is delivered by a multi-disciplinary team consisting of specialist nurses, pharmacists, peer support worker, dietician, and clinicians with specialist interest in inclusion health (an umbrella term describing people who experience extreme social exclusion and multiple barriers to accessing healthcare)^12,13^.

### Data collection and definitions

We extracted a list of all adults (18 years and older) living with chronic hepatitis B who had a transient elastography with controlled attenuation parameter (CAP) score from the electronic patient record (EPR) between 1st May 2018 (when CAP was introduced to the clinic) and 28th February 2024 (date of data extraction). CAP is a quantitative ultrasound-based measure of liver steatosis, assessed using a module built into the Fibroscan^TM^. Overall data collection was completed by August 2024. We defined chronic hepatitis B on the basis of the clinic physician evaluation, incorporating a combination of clinical history and blood tests. We assigned the most recent CAP score test date as the index date. We only included participants who attended at least one clinic appointment and had monitoring blood tests within 2 years of their CAP score.

Participants were excluded if they had antibodies to hepatitis C or delta viruses (i.e. past or present hepatitis C virus (HCV), or hepatitis D virus (HDV) infection) or HIV. HCV is steatogenic and HIV increases cardiovascular risk, therefore are confounding variables in the relationship between CHB and MASLD^14,15^. The relationship between HDV and MASLD is unknown, however HDV is known to affect CHB outcomes^16^. Since there were only 10 people with positive anti-HDV antibody tests, too small for subgroup analysis, we excluded this group from the final analysis.

Detailed information on the variables collected, definitions and methods of collection can be seen in **Table 1**. In brief, we collected data on demographics (age, sex, ethnicity), cardiometabolic factors (diabetes, obesity, dyslipidaemia, hypertension), viral parameters (hepatitis B e-Antigen (HBeAg), viral load (HBV VL), quantitative surface antigen (qHBsAg), nucleot(s)ide analogue (NA) treatment) and liver disease (liver stiffness measured by Fibroscan^TM^) alanine transaminase (ALT), aspartate amino-transferase (AST), gamma-glutamyl transferase (GGT)) from the electronic patient record (EPR).

**Table 1:**
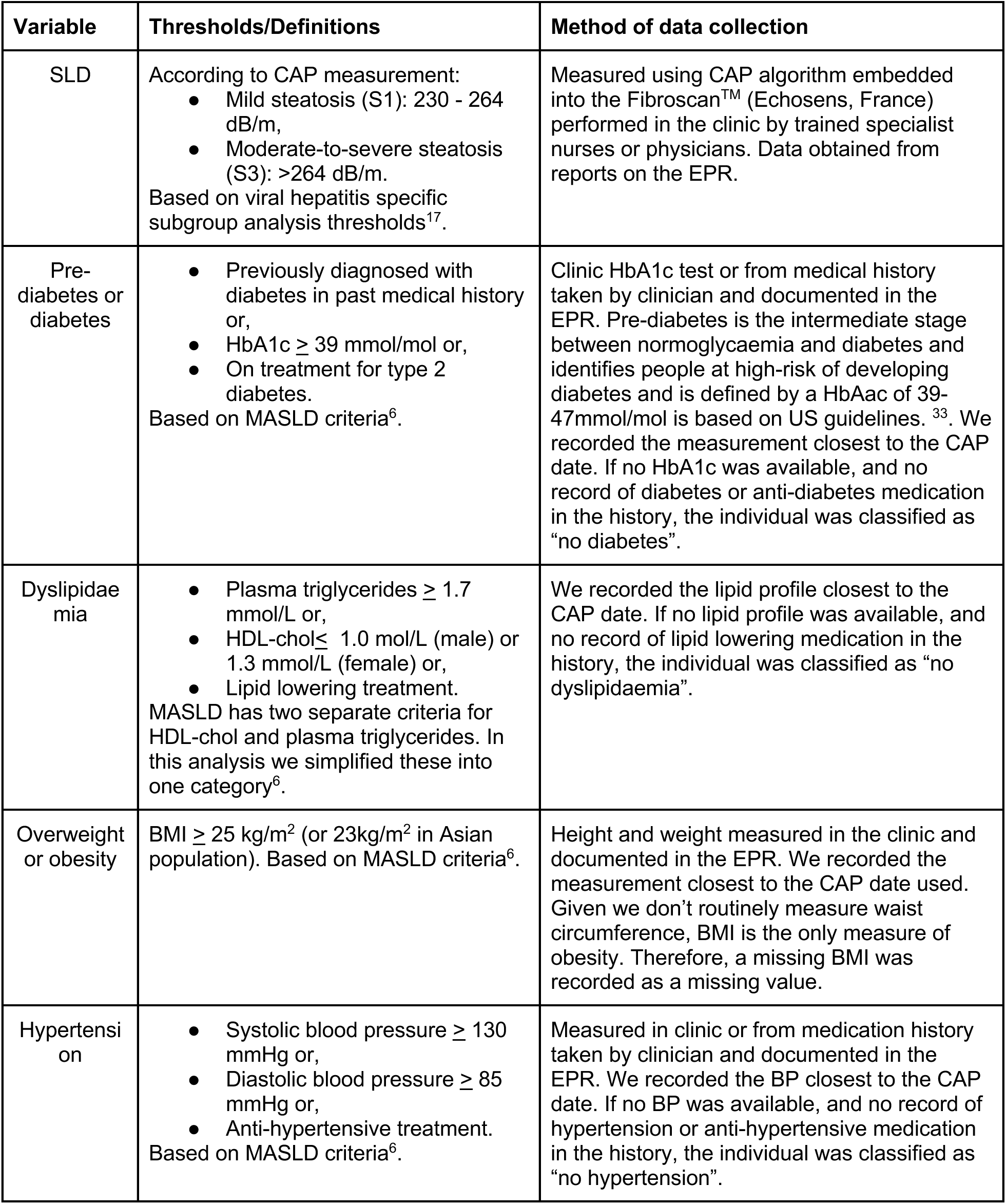
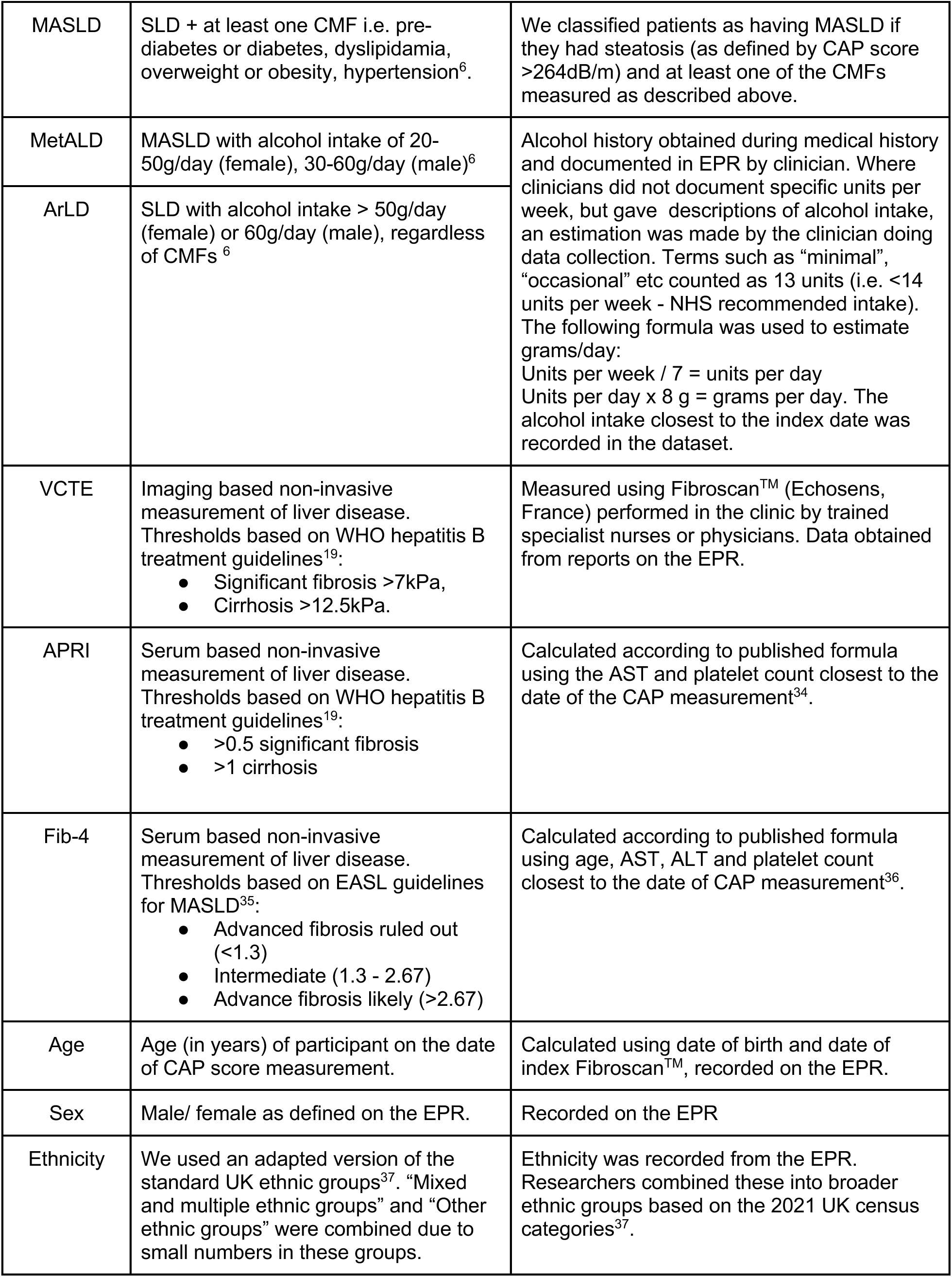
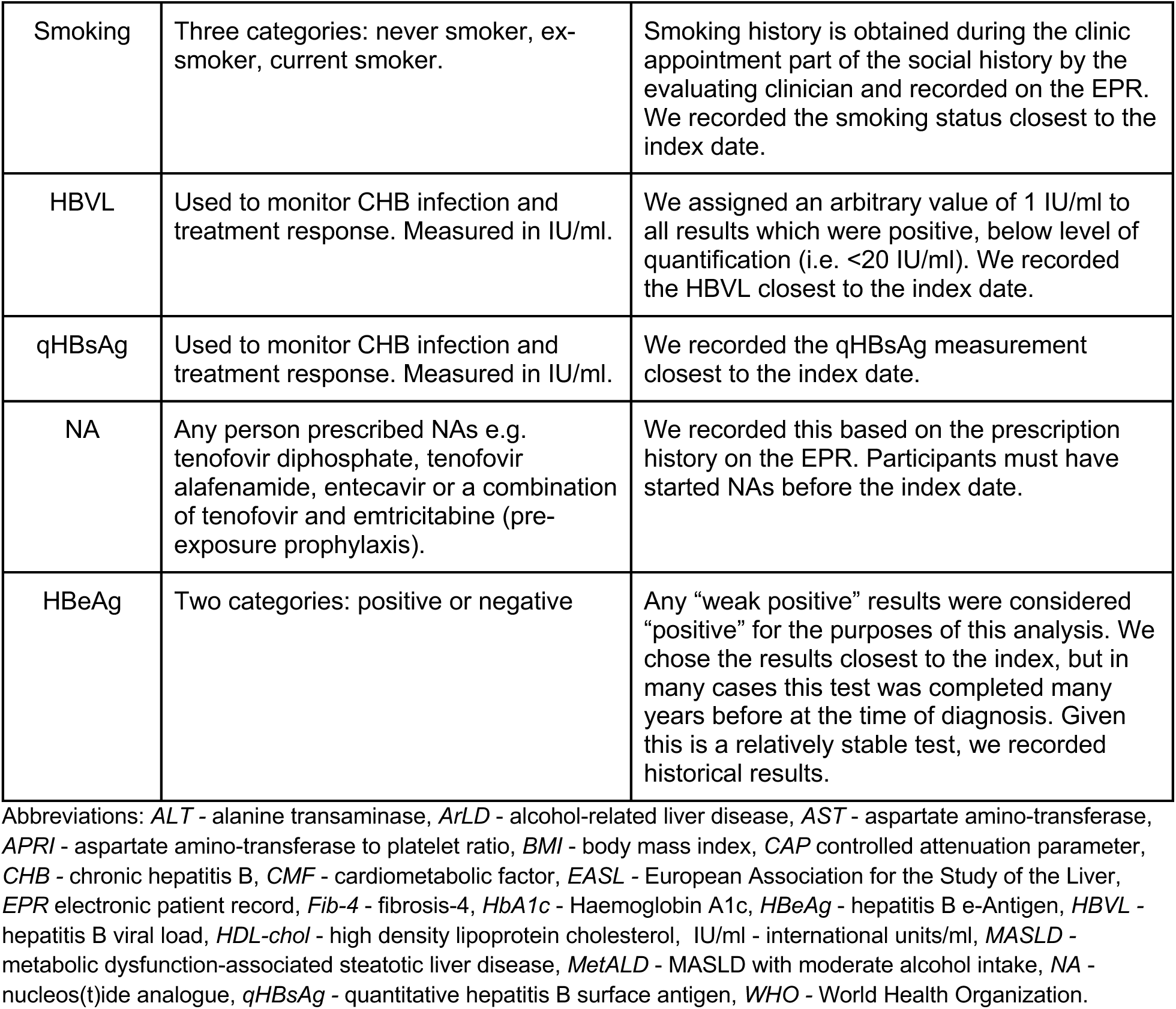
Definitions, thresholds and methods of data collection of key variables included in the study to investigate the relationship between metabolic dysfunction-associated steatotic liver disease (MASLD) and chronic hepatitis B (CHB) in people attending a London viral hepatitis clinic.

| Variable | Thresholds/Definitions | Method of data collection |
| --- | --- | --- |
| SLD | According to CAP measurement:<br>Mild steatosis (S1): 230 - 264 dB/m,<br>Moderate-to-severe steatosis (S3): >264 dB/m.<br>Based on viral hepatitis specific subgroup analysis thresholds<sup>17</sup>. | Measured using CAP algorithm embedded into the Fibroscan™ (Echosens, France) performed in the clinic by trained specialist nurses or physicians. Data obtained from reports on the EPR. |
| Pre-diabetes or diabetes | Previously diagnosed with diabetes in past medical history or,<br>HbA1c ≥ 39 mmol/mol or,<br>On treatment for type 2 diabetes.<br>Based on MASLD criteria<sup>6</sup>. | Clinic HbA1c test or from medical history taken by clinician and documented in the EPR. Pre-diabetes is the intermediate stage between normoglycaemia and diabetes and identifies people at high-risk of developing diabetes and is defined by a HbAac of 39-47mmol/mol is based on US guidelines.<sup>33</sup>. We recorded the measurement closest to the CAP date. If no HbA1c was available, and no record of diabetes or anti-diabetes medication in the history, the individual was classified as “no diabetes”. |
| Dyslipidaemia | Plasma triglycerides ≥ 1.7 mmol/L or,<br>HDL-cholesterol ≤ 1.0 mmol/L (male) or 1.3 mmol/L (female) or,<br>Lipid lowering treatment.<br>MASLD has two separate criteria for HDL-cholesterol and plasma triglycerides. In this analysis we simplified these into one category<sup>6</sup>. | We recorded the lipid profile closest to the CAP date. If no lipid profile was available, and no record of lipid lowering medication in the history, the individual was classified as “no dyslipidaemia”. |
| Overweight or obesity | BMI ≥ 25 kg/m<sup>2</sup> (or 23kg/m<sup>2</sup> in Asian population). Based on MASLD criteria<sup>6</sup>. | Height and weight measured in the clinic and documented in the EPR. We recorded the measurement closest to the CAP date used. Given we don't routinely measure waist circumference, BMI is the only measure of obesity. Therefore, a missing BMI was recorded as a missing value. |
| Hypertension | Systolic blood pressure ≥ 130 mmHg or,<br>Diastolic blood pressure ≥ 85 mmHg or,<br>Anti-hypertensive treatment.<br>Based on MASLD criteria<sup>6</sup>. | Measured in clinic or from medication history taken by clinician and documented in the EPR. We recorded the BP closest to the CAP date. If no BP was available, and no record of hypertension or anti-hypertensive medication in the history, the individual was classified as “no hypertension”. |
| MASLD | SLD + at least one CMF i.e. pre-diabetes or diabetes, dyslipidamia, overweight or obesity, hypertension<sup>6</sup>. | We classified patients as having MASLD if they had steatosis (as defined by CAP score >264dB/m) and at least one of the CMFs measured as described above. |
| MetALD | MASLD with alcohol intake of 20-50g/day (female), 30-60g/day (male)<sup>6</sup> | Alcohol history obtained during medical history and documented in EPR by clinician. Where clinicians did not document specific units per week, but gave descriptions of alcohol intake, an estimation was made by the clinician doing data collection. Terms such as “minimal”, “occasional” etc counted as 13 units (i.e. <14 units per week - NHS recommended intake). The following formula was used to estimate grams/day:<br>Units per week / 7 = units per day<br>Units per day x 8 g = grams per day. The alcohol intake closest to the index date was recorded in the dataset. |
| ArLD | SLD with alcohol intake > 50g/day (female) or 60g/day (male), regardless of CMFs<sup>6</sup> |  |
| VCTE | Imaging based non-invasive measurement of liver disease. Thresholds based on WHO hepatitis B treatment guidelines<sup>19</sup>:<br>• Significant fibrosis >7kPa,<br>• Cirrhosis >12.5kPa. | Measured using Fibroscan<sup>TM</sup> (Echosens, France) performed in the clinic by trained specialist nurses or physicians. Data obtained from reports on the EPR. |
| APRI | Serum based non-invasive measurement of liver disease. Thresholds based on WHO hepatitis B treatment guidelines<sup>19</sup>:<br>• >0.5 significant fibrosis<br>• >1 cirrhosis | Calculated according to published formula using the AST and platelet count closest to the date of the CAP measurement<sup>34</sup>. |
| Fib-4 | Serum based non-invasive measurement of liver disease. Thresholds based on EASL guidelines for MASLD<sup>35</sup>:<br>• Advanced fibrosis ruled out (<1.3)<br>• Intermediate (1.3 - 2.67)<br>• Advance fibrosis likely (>2.67) | Calculated according to published formula using age, AST, ALT and platelet count closest to the date of CAP measurement<sup>36</sup>. |
| Age | Age (in years) of participant on the date of CAP score measurement. | Calculated using date of birth and date of index Fibroscan<sup>TM</sup>, recorded on the EPR. |
| Sex | Male/ female as defined on the EPR. | Recorded on the EPR |
| Ethnicity | We used an adapted version of the standard UK ethnic groups<sup>37</sup>. “Mixed and multiple ethnic groups” and “Other ethnic groups” were combined due to small numbers in these groups. | Ethnicity was recorded from the EPR. Researchers combined these into broader ethnic groups based on the 2021 UK census categories<sup>37</sup>. |
| Smoking | Three categories: never smoker, ex-smoker, current smoker. | Smoking history is obtained during the clinic appointment part of the social history by the evaluating clinician and recorded on the EPR. We recorded the smoking status closest to the index date. |
| HBVL | Used to monitor CHB infection and treatment response. Measured in IU/ml. | We assigned an arbitrary value of 1 IU/ml to all results which were positive, below level of quantification (i.e. <20 IU/ml). We recorded the HBVL closest to the index date. |
| qHBsAg | Used to monitor CHB infection and treatment response. Measured in IU/ml. | We recorded the qHBsAg measurement closest to the index date. |
| NA | Any person prescribed NAs e.g. tenofovir diphosphate, tenofovir alafenamide, entecavir or a combination of tenofovir and emtricitabine (pre-exposure prophylaxis). | We recorded this based on the prescription history on the EPR. Participants must have started NAs before the index date. |
| HBeAg | Two categories: positive or negative | Any “weak positive” results were considered “positive” for the purposes of this analysis. We chose the results closest to the index, but in many cases this test was completed many years before at the time of diagnosis. Given this is a relatively stable test, we recorded historical results. |

Abbreviations: *ALT* - alanine transaminase, *ArLD* - alcohol-related liver disease, *AST* - aspartate amino-transferase, *APRI* - aspartate amino-transferase to platelet ratio, *BMI* - body mass index, *CAP* controlled attenuation parameter, *CHB* - chronic hepatitis B, *CMF* - cardiometabolic factor, *EASL* - European Association for the Study of the Liver, *EPR* electronic patient record, *Fib-4* - fibrosis-4, *HbA1c* - Haemoglobin A1c, *HBeAg* - hepatitis B e-Antigen, *HBVL* - hepatitis B viral load, *HDL-cho* - high density lipoprotein cholesterol, *IU/ml* - international units/ml, *MASLD* - metabolic dysfunction-associated steatotic liver disease, *MetALD* - MASLD with moderate alcohol intake, *NA* - nucleos(t)ide analogue, *qHBsAg* - quantitative hepatitis B surface antigen, *WHO* - World Health Organization.

There is variation regarding optimal CAP steatosis thresholds in the literature and we opted to use thresholds to balance sensitivity and specificity, using the moderate-to-severe (S2/3) steatosis (CAP > 264 dB/m) vs no/mild steatosis (CAP<264 dB/m), which we will henceforth refer to as “MASLD” and “no MASLD”, for all comparative analyses in this study^17^.

### Missing data

Since dyslipidaemia, hypertension and prediabetes/diabetes are composite variables with broad inclusion criteria according to the MASLD definition (**Table 1**), we took a pragmatic approach and if an individual did not have a recorded lipid profile and had no record of lipid-lowering medications we classified as “no dyslipidaemia”, The same approach applied to hypertension and prediabetes/diabetes. Since overweight/obesity is based only on BMI (waist circumference is not routinely measured in clinic), we counted those without a BMI measurement as missing data (i.e. unable to classify them as being overweight/obese or normal/underweight) and applied an available case approach to analysis. Missing data are detailed in the footnote of **Table 2**.

**Table 2:**
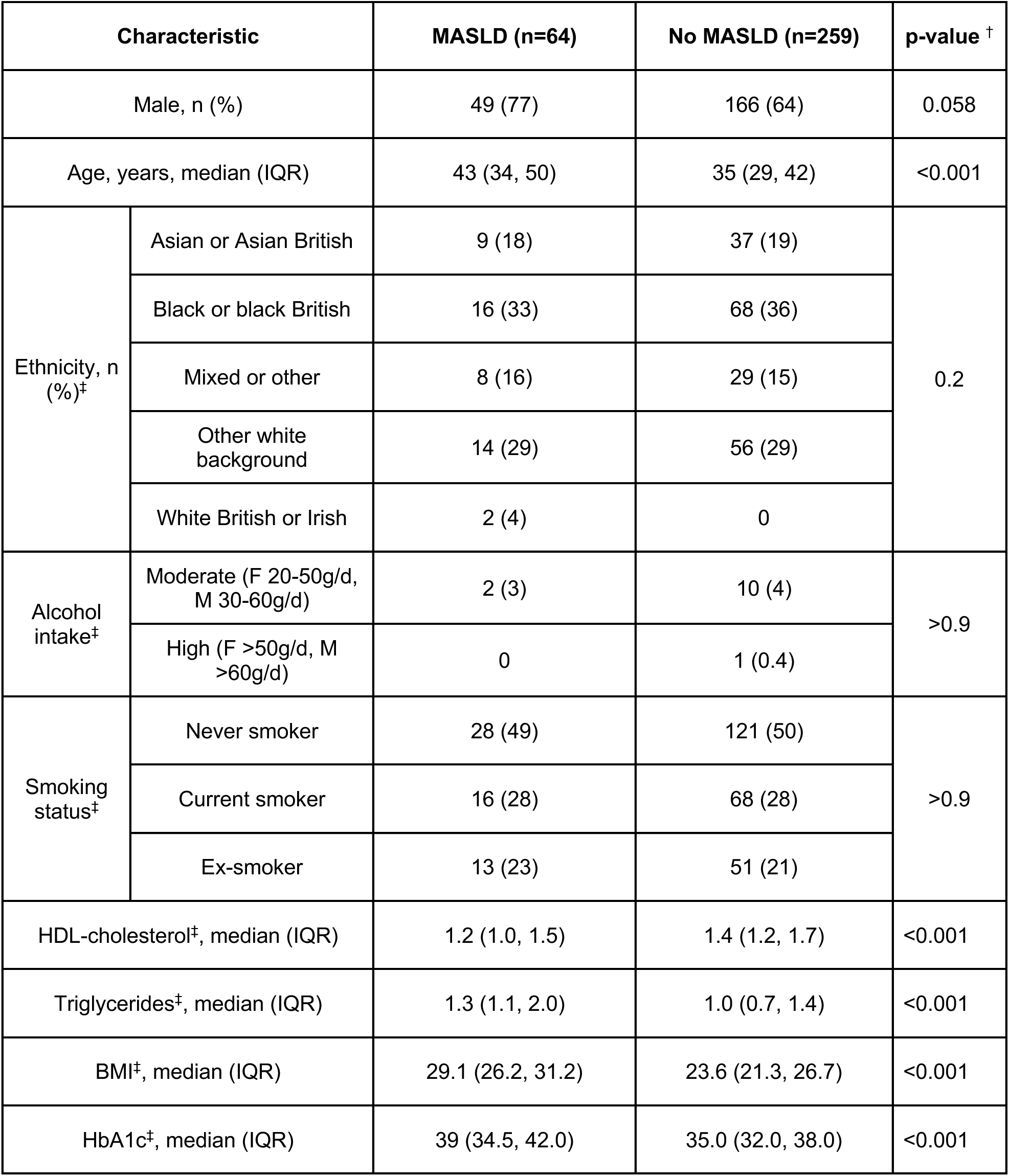
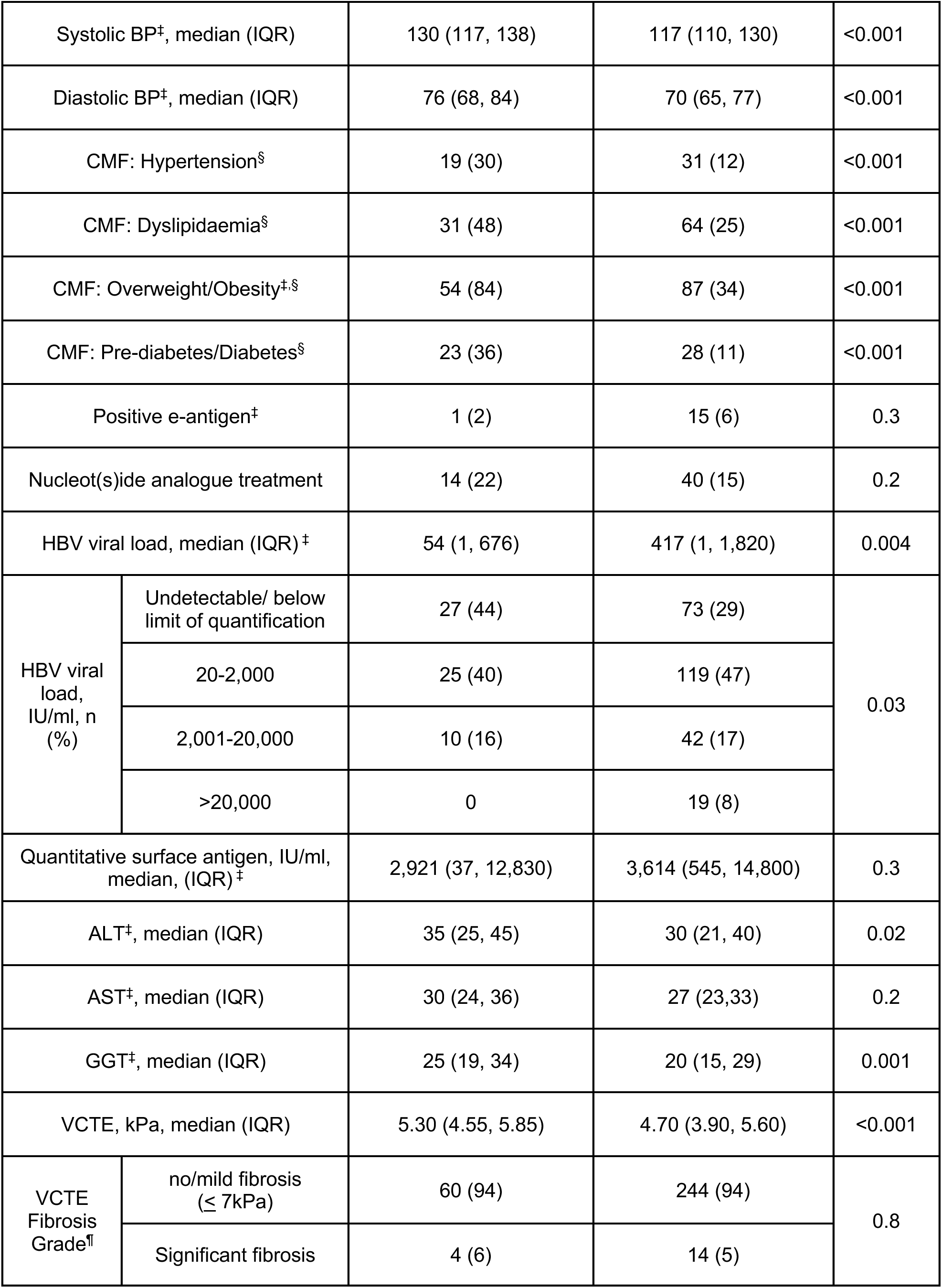
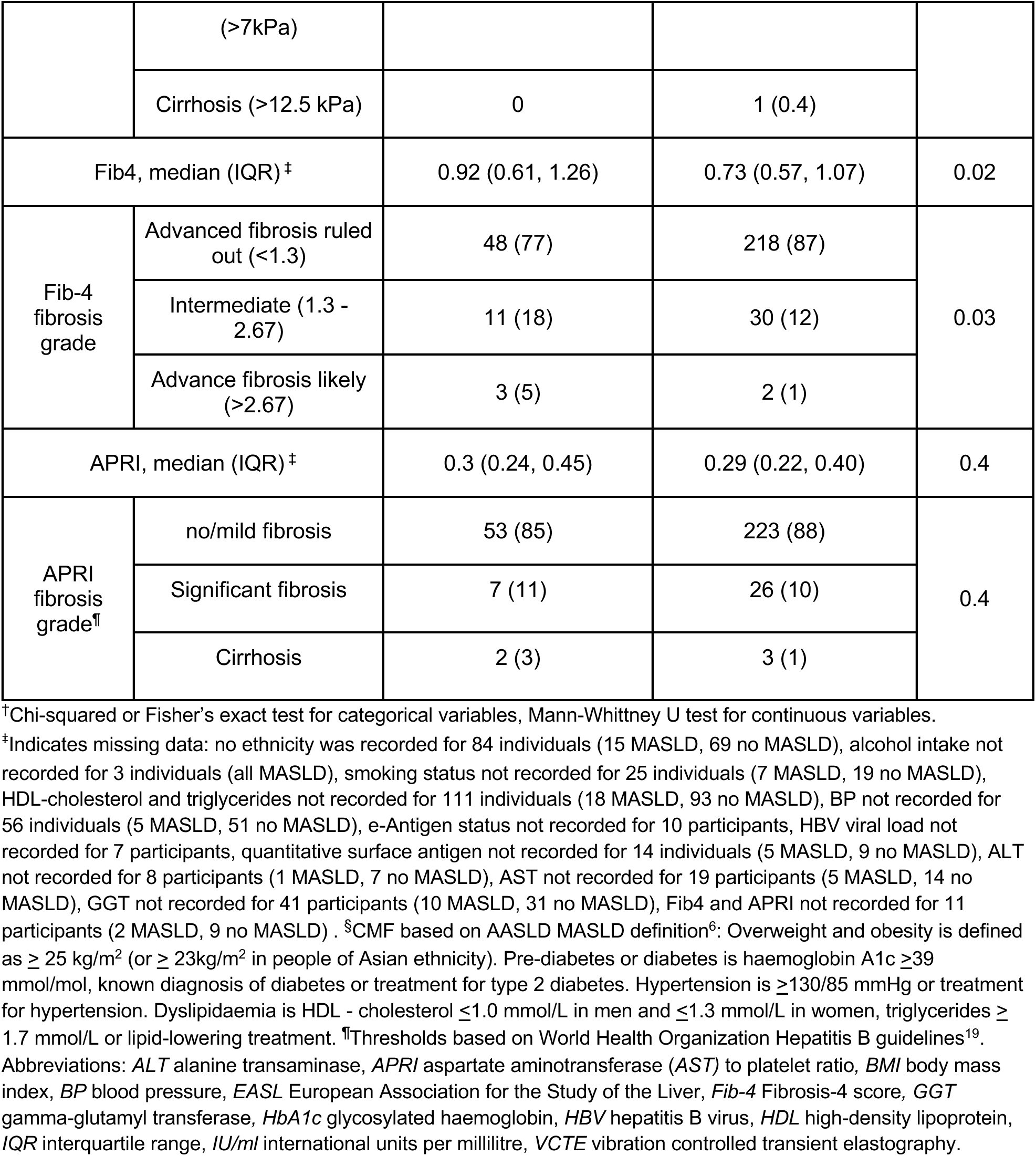
Demographic characteristics, viral parameters and liver disease outcomes among people living with chronic hepatitis B (CHB) with and without metabolic dysfunction-associated steatotic liver disease (MASLD). Thresholds for MASLD based on viral hepatitis specific controlled attenuation parameter (CAP) thresholds: CAP >230 dB/m - mild steatosis, CAP>264 dB/m - moderate/severe steatosis^17^.

### Data analysis

We summarised continuous variables using median and interquartile range, and categorical variables using total number and percentage. We compared demographic, cardiometabolic, viral and liver disease parameters according to MASLD status, using Mann-Whitney-U and Kruskal-wallis tests for continuous variables and Chi-squared (or Fisher’s exact test where expected counts were less than 5) for categorical variables. For further analyses, we log10 transformed HBV viral load to reduce the right-skewness of the distribution and to improve the fit of linear regression models. We added 1 to any undetectable viral load, since it is not possible to log transform 0. We analysed the data using R Studio (v4.5.1).

We investigated the impact of MASLD on liver stiffness using a multivariable linear regression model. Using a directed acyclic graph (DAG) we identified a minimum adjustment set of sex, age, ethnicity, alcohol, smoking status and NA treatment (**Suppl Figure 1**). As any relationship between MASLD and HBV viral load is hypothetical, our DAG does not identify this as a confounding variable in the minimum adjustment set, however, given the known relationship between HBV viral load and liver disease we included HBV viral load in our model^18^.

We further investigated the relationship between metabolic disease and HBV viral load. We compared log_10_(HBV viral load) across the following categories:

- presence or absence of each CMF (prediabetes/diabetes, dyslipidaemia, overweight/obesity, hypertension) using Mann-Whitney U tests,
- steatosis grade, number of CMFs using Kruskal-Wallis test (comparison across multiple categories).

We used linear regression to investigate the relationship between MASLD and HBV viral load. Using a DAG to map the hypothetical relationship between MASLD and HBV viral load, we identified a minimum sufficient adjustment set, which included age, ethnicity, nucleos(t)ide analogue (NA) treatment and sex (**Suppl Figure 2**). There is no current evidence to suggest hepatitis B e-antigen (HBeAg) status is associated with MASLD, therefore we have not included this relationship in the DAG. However, HBeAg is associated with HBV viral load, therefore we opted to include it in the model (as a “competing exposure”) to improve precision of the estimate.

For both linear regression models, a global p value for ethnicity (a categorical variable with multiple levels) was derived from an F-test comparing the full model to a reduced model excluding ethnicity. In addition, for both models, a sensitivity analysis was performed considering only the untreated population, given the effect of antiviral treatment on HBV viral load suppression and improving/preventing liver disease.

### Ethical Approval and Governance

This study received NHS Health Research Authority approval (reference 25/HRA/4006) and was undertaken in keeping with the Declaration of Helsinki.

## Results

### Cohort description

Between May 2018 and February 2024, 323 people living with CHB were eligible to be included in this study (**Figure 1**). Sixty-seven percent (215/323) were male with a median age of 36 years (interquartile range 30, 44 years). People of black ethnicity were the largest group (84/239, 35%), followed by white (other than British/Irish) (29%, 70/239) (**Table 2**). Sixty-four percent (207/323) had at least one CMF, and the most common metabolic comorbidity was overweight/obesity (141/279, 50%). In terms of steatotic liver disease, 18% (59/323) were categorised as mild (CAP 230-264 dB/m) and 23% (73/323) as moderate/severe (CAP >264 dB/m). MASLD (CAP >264 dB/m + at least one CMF) was present in 19% (63/323) and MASLD with moderate alcohol intake (MetALD) was present in 0.6% (2/323) of the total population (**Figure 2**, **Table 2**).

**Figure 1:**
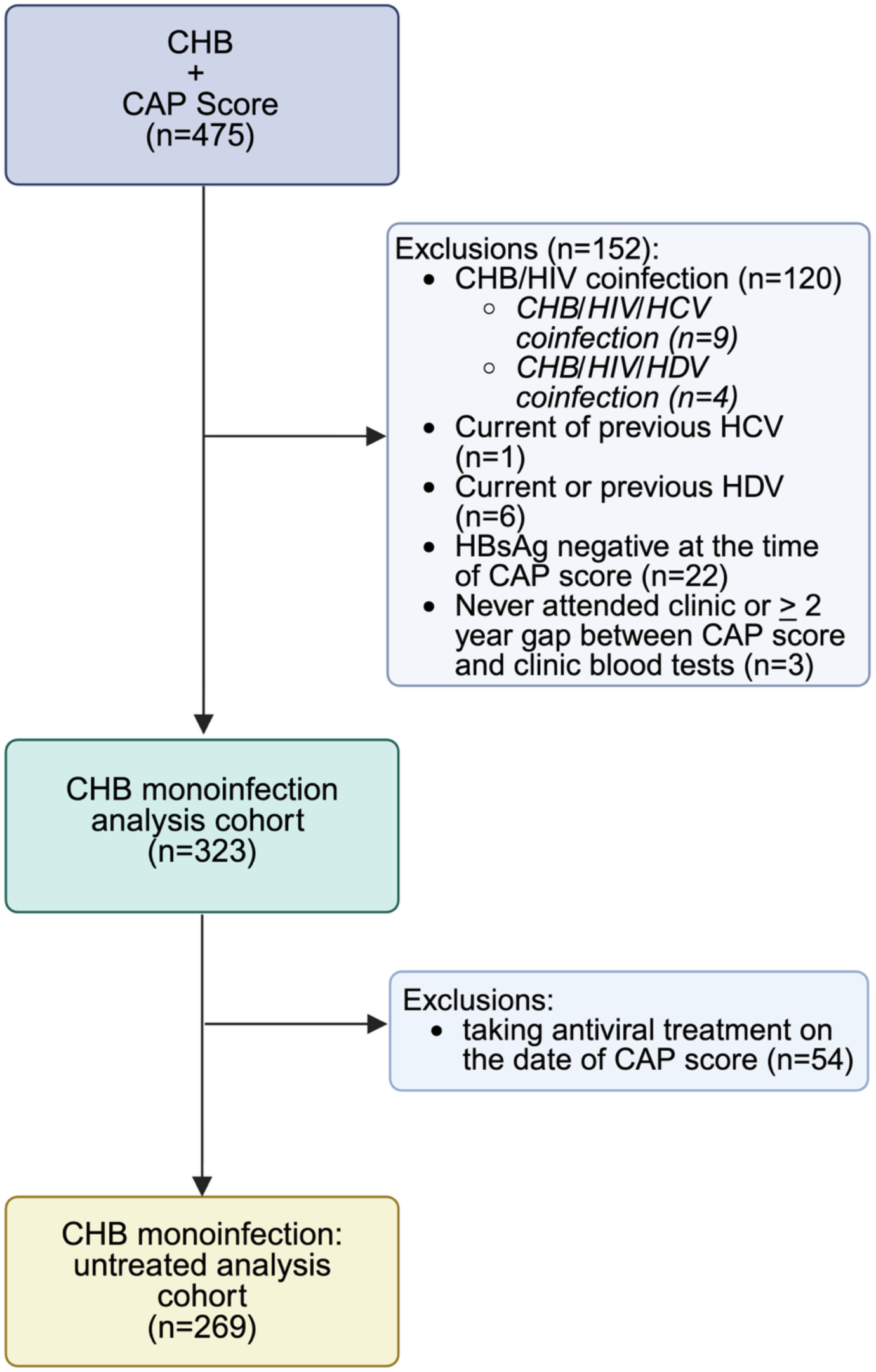
Study flow diagram showing the selection of adults living with chronic hepatitis B (CHB) attending a central London clinic for a study investigating the relationship between CHB and metabolic dysfunction-associated steatotic liver disease (MASLD). Abbreviations: *CAP* - controlled attenuation parameter, *CHB -* chronic hepatitis B, *HCV* - hepatitis C virus, *HDV* - hepatitis D virus, *HIV* - human immunodeficiency virus.

**Figure 2:**
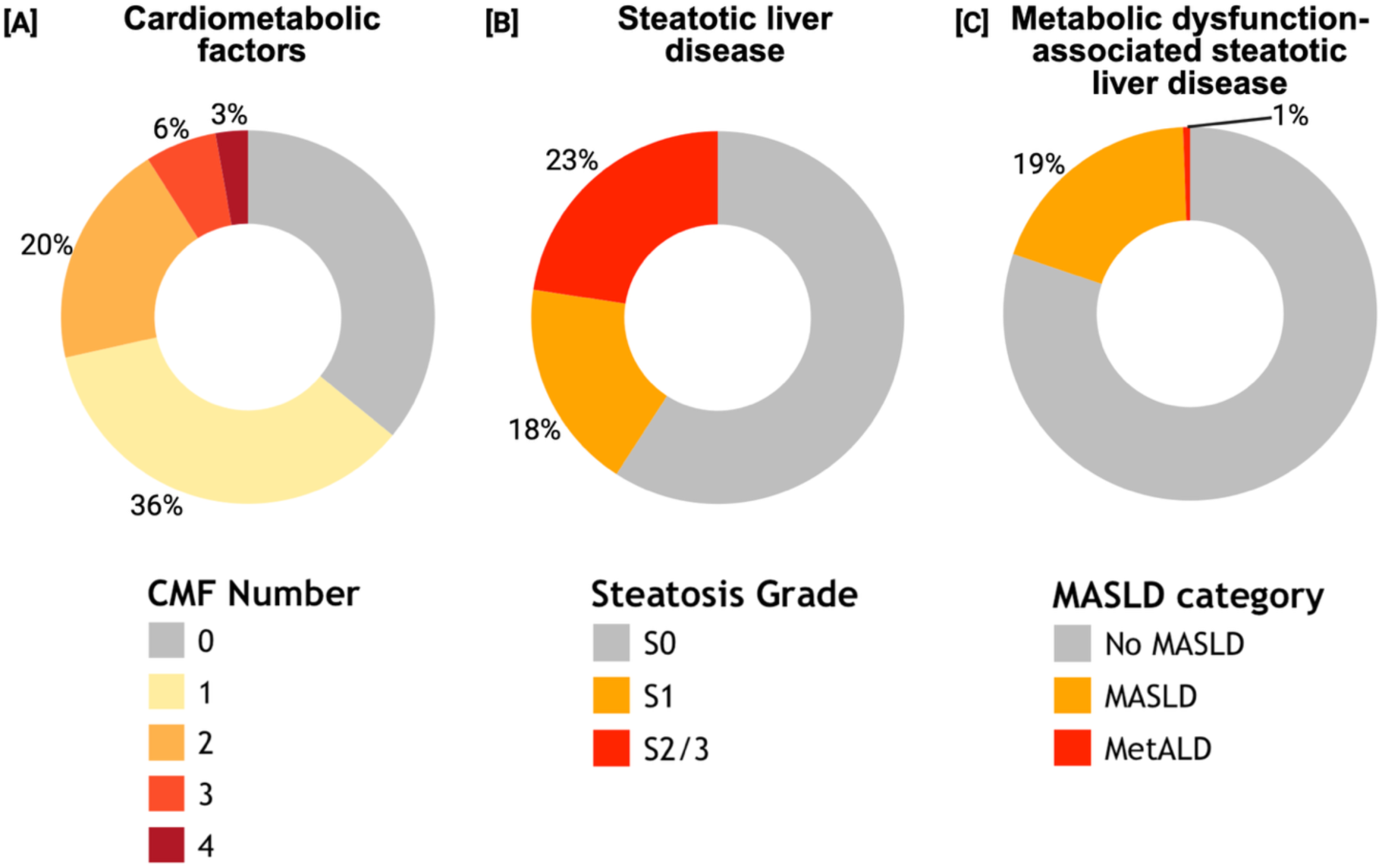
Cardiometabolic factors (CMFs, hypertension, overweight and obesity, prediabetes and type 2 diabetes, dyslipidaemia), steatotic liver disease severity and metabolic dysfunction-associated steatotic liver disease (MASLD) among people living with chronic hepatitis B attending a central London clinic. Donut plots illustrating **[A]** Number of CMFs present in each individual, **[B]** steatotic liver disease (SLD) severity; S1 (mild) CAP >230 dB/m, S2/3 (moderate/severe) CAP >264 dB/m, **[C]** MASLD in the total population; Abbreviations: MetALD - MASLD + moderate alcohol intake (20-50g/day females, 30-60g/day males), HBV - hepatitis B virus, HIV - human immunodeficiency virus.

### Characteristics of the population living with MASLD

Compared to people with no MASLD, people with MASLD were significantly older (median age (IQR): 43 (34, 50) vs 35 (29, 42) years, p<0.001), but there was no significant difference in sex or ethnicity (**Table 2**). There was significantly more liver disease in the MASLD group, which had a higher median ALT (35 (25, 45) vs 30 (21, 40) IU/L, p=0.02), gamma-glutamyl transaminase (25 (19, 34) vs 20 (15, 29) IU/L, p=0.001), liver stiffness score (5.3 (4.6, 5.9) vs 4.7 (3.8, 5.6) kPa, p <0.001), and more people with significant fibrosis (4.8% vs 0.8%, p=0.03) according to their Fib-4 score (**Table 1**). However, there were no significant differences in aspartate aminotransferase (AST), aspartate aminotransferase-to-platelet ratio index (APRI) score or liver stiffness measurement when categorised according WHO thresholds for significant fibrosis (>7kPa) and cirrhosis (>12.5kPa)^19^.

### Relationship between MASLD and liver disease in people living with CHB

People with MASLD had a significantly higher liver stiffness than those with no MASLD in a multivariable linear regression model (β = 0.48kPa, 95% CI 0.06 to 0.90, p=0.03). Ethnicity was significantly associated with liver stiffness, and people of Black (β = 0.79kPa, 95% CI 0.32 to 1.27, p=0.001) and mixed/other ethnicity (β = 1.05kPa, 95% CI 0.50 to 0.1.63, p<0.001) had significantly higher liver stiffness scores compared to people of Asian ethnicity (**Table 3**)

**Table 3:**
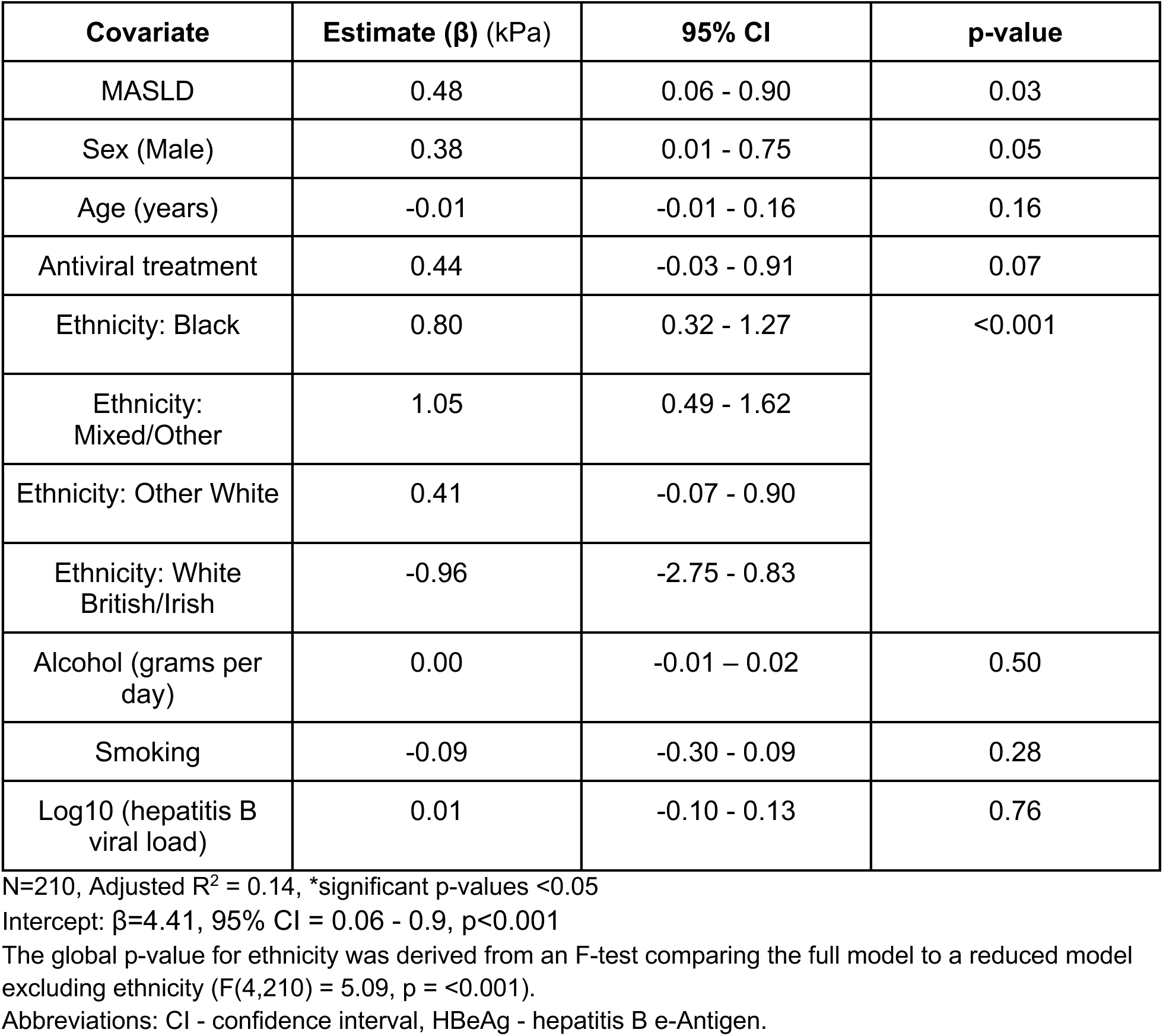
Results of a multiple linear regression model to see how metabolic dysfunction-associated steatotic liver disease (MASLD) and other factors are jointly associated with liver stiffness (outcome) in people living with chronic hepatitis B. MASLD (controlled attenuation parameter >264 dB/m and at least one cardiometabolic factor). Reference ethnicity = Asian, reference sex = female.

| **Covariate** | **Estimate ($\beta$) (kPa)** | **95% CI** | **p-value** |
| --- | --- | --- | --- |
| MASLD | 0.48 | 0.06 - 0.90 | 0.03 |
| Sex (Male) | 0.38 | 0.01 - 0.75 | 0.05 |
| Age (years) | -0.01 | -0.01 - 0.16 | 0.16 |
| Antiviral treatment | 0.44 | -0.03 - 0.91 | 0.07 |
| Ethnicity: Black | 0.80 | 0.32 - 1.27 | <0.001 |
| Ethnicity: Mixed/Other | 1.05 | 0.49 - 1.62 |  |
| Ethnicity: Other White | 0.41 | -0.07 - 0.90 |  |
| Ethnicity: White British/Irish | -0.96 | -2.75 - 0.83 |  |
| Alcohol (grams per day) | 0.00 | -0.01 – 0.02 | 0.50 |
| Smoking | -0.09 | -0.30 - 0.09 | 0.28 |
| Log10 (hepatitis B viral load) | 0.01 | -0.10 - 0.13 | 0.76 |

N=210, Adjusted $R^2$ = 0.14, \*significant p-values <0.05
Intercept: $\beta$ =4.41, 95% CI = 0.06 - 0.9, p<0.001
The global p-value for ethnicity was derived from an F-test comparing the full model to a reduced model excluding ethnicity ( $F(4,210) = 5.09$ , $p = <0.001$ ).
Abbreviations: CI - confidence interval, HBeAg - hepatitis B e-Antigen.

### Relationship between MASLD and CHB

HBV viral load was significantly lower in people with MASLD (median HBV VL 54 (<20, 676) vs 417 (<20, 820) IU/ml, p=0.004), despite there being no significant difference in HBeAg positivity (1.6% vs 5.7%, p=0.2) or people on antiviral treatment (22% vs 15%, p=0.2) (**Table 2**). There was an inverse trend, and significant overall association, between HBV viral load and number of CMF per person, steatosis grade, and number of CMF per person with MASLD (**Figure 3**). HBV viral load was significantly lower in people living with, compared to people without, diabetes or pre-diabetes (median log10 viral load 1.56 vs 2.41, p = 0.02), overweight or obesity (median log10 viral load 2.28 vs 2.74, p = 0.008) and dyslipidaemia (median log10 viral load 2.06 vs 2.61, p = 0.006), but there was no significant difference in median viral load according to hypertension status (median log10 viral load 2.40 vs 2.49, p = 0.78) (**Figure 3**).

**Figure 3:**
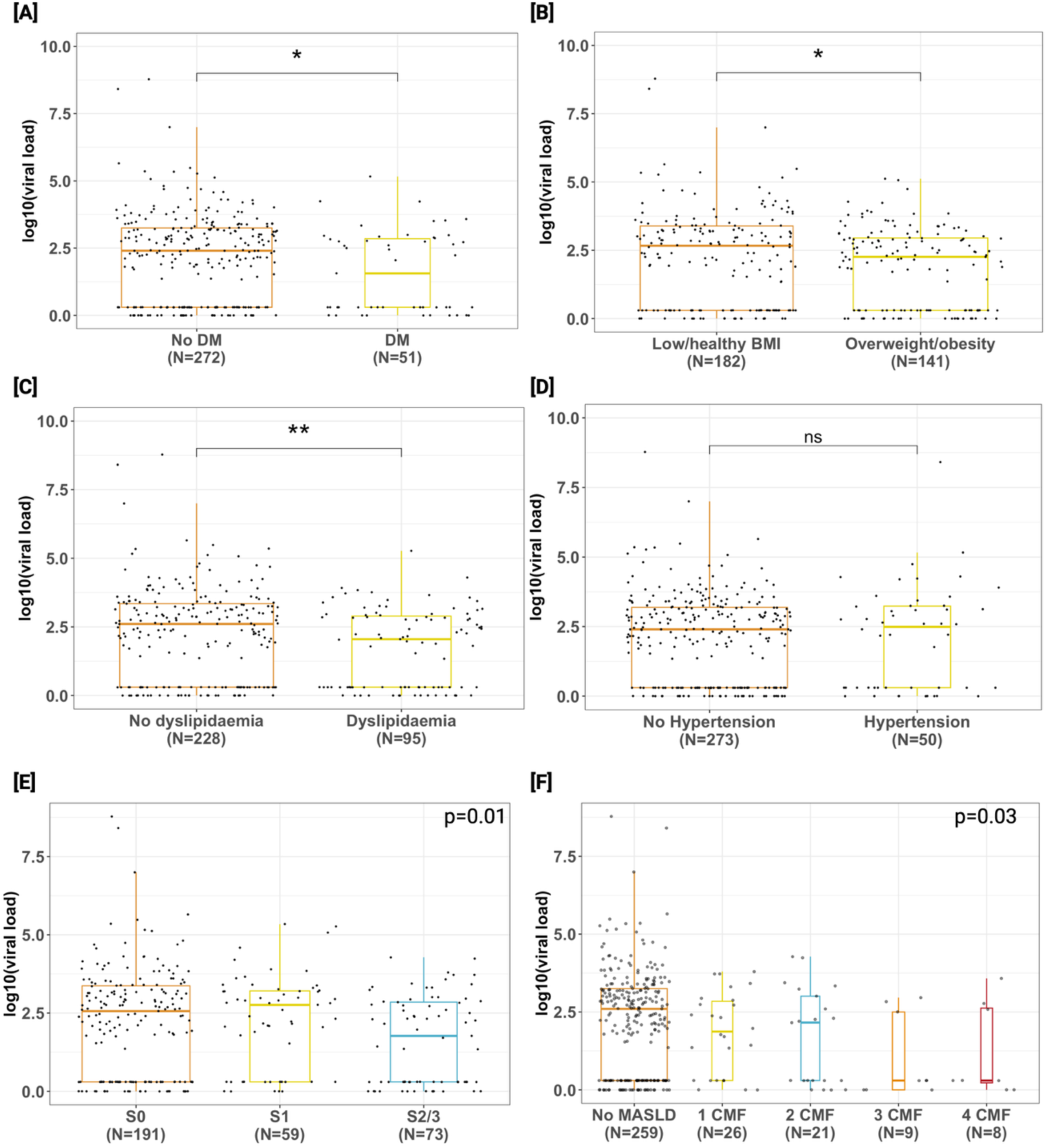
Comparison of hepatitis B virus (HBV) viral load distributions across [A] type 2 diabetes and pre diabetes, [B] overweight/obesity, [C] dyslipidaemia, [D] hypertension, [E] steatotic liver disease (SLD) grade and [F] number of cardiometabolic factors (CMF) per person with moderate-severe metabolic dysfunction-associated steatotic liver disease (MASLD). CMF are defined according to the European Association for the Study of the Liver (EASL) metabolic dysfunction associated steatotic liver disease (MASLD) guidelines^35^. Mann-Whitney U test was used to compare viral load between two groups. Kruskal-wallis was used to compare viral load between more than 2 groups. * = p<0.05, ** = p<0.01. Abbreviations: DM - pre diabetes and type 2 diabetes mellitus, ns - not significant. S0 = no SLD, S1 = mild SLD (controlled attenuation parameter (CAP) >230 dB/m), S2/3 = mod-severe SLD (CAP >264 dB/m).

In a multivariable linear regression model investigating the association between MASLD and HBV viral load, antiviral treatment was independently associated with lower HBV viral load (β = -1.97, 95% CI -2.42 to -1.53, p=<0.001), while HBeAg positivity was associated with a higher viral load (β = 1.13, 95% CI 0.30 to 1.96, p=0.009). There was no association between MASLD and HBV viral load after adjustment for confounders (age, sex, antiviral treatment, ethnicity, (β = -0.29 95% CI -0.73 to 0.15, p=0.2, **Suppl Table 2**).

### Sensitivity analyses

The effect of MASLD on liver disease was more pronounced in people not taking antiviral treatment (β = 0.57kPa 95% CI 0.12 – 1.03, p=0.01) (**Suppl Table 3**). We performed a sensitivity analysis to investigate the effect of MASLD on HBV viral load in people not taking antiviral treatment, given the confounding effect of this variable. The association with MASLD was not significant and HBeAg remained the most important predictor of viral load (β = 1.33, 95% CI 0.58 to 2.29, p =0.02) (**Suppl Table 4**). Further sensitivity analyses of the HBeAg negative population were not reported, since the model’s explained variance was low (<3%), limiting interpretation.

## Discussion

### Key Findings

We found that metabolic comorbidity is common in people living with CHB attending a diverse central London clinic. Of clinical importance, MASLD is associated with liver disease outcomes, and was significantly associated with markers of liver disease, including liver stiffness in this study population. An interesting, inverse association between HBV viral load and MASLD was observed in univariable analysis, but this was not maintained after adjustment, suggesting no independent relationship or lack of study power to detect a true effect.

### Results in context

Approximately 30% of the global population is affected by MASLD^3^. We are in the midst of a global obesity epidemic, since prevalence doubled between 1990 and 2021 and by 2050, an estimated 1.9 billion adults will be living with overweight and obesity^20^. The prevalence of MASLD in people living with CHB is estimated to reflect that of the general population (35%, 95% CI 32-38%)^21^. Fewer people have MASLD (20%) in this cohort, but CMFs are common; 64% have at least one CMF, most frequently overweight or obesity.

While our study reports an association between MASLD and increased liver stiffness, ALT, GGT and Fib-4 score, other published studies do not demonstrate a consistent association between MASLD and liver disease outcomes. Some report an increased risk of cirrhosis and HCC in people living with both MASLD and CHB compared to CHB alone, while others suggest no association, or even a decreased risk^9^. The reasons for this heterogeneity are not clear, but may include differences in population, study design, MASLD diagnosis and statistical methods^22,23^.

One important source of variation is method and threshold for steatosis diagnosis. In our study, we applied a CAP threshold of 264 dB/m, based on a viral hepatitis specific individual patient data meta-analysis^17^. Given the wide range of thresholds used in the literature, we selected this moderate-severe cut-off (264 dB/m) rather than using the lower threshold suggested in the study for mild steatosis (230dB/m). Such variation in diagnostic thresholds is likely to contribute to inconsistencies in findings across studies.

We did not observe an association between MASLD and liver stiffness categories based on WHO thresholds. This likely reflects the small absolute difference between groups (approximately 0.5 kPa higher in the MASLD group), together with the relatively low prevalence of advanced liver disease within our cohort. In addition, unlike ALT and GGT, AST was not significantly increased in the MASLD group, which is a typical MASLD LFT pattern^24^. AST levels typically rise with increasing alcohol intake and advancing liver disease, particularly with progression to fibrosis.

Therefore, the absence of a difference in AST in this cohort may reflect both the relatively low alcohol exposure and the overall low burden of advanced liver disease in this population. ^24^.

Our observation of an inverse relationship between CHB, CMFs and MASLD is supported by several studies in the literature. MASLD, and obesity, have been associated with increased spontaneous HBsAg seroclearance in untreated CHB, with one longitudinal study observing a dose-dependent increase in probability of seroclearance of MASLD plus increasing number of CMFs ^7,9,25–27^. In addition, several studies have reported inverse associations between HBV viral load and MASLD, obesity and triglycerides^28–30^. Caution should be used in interpreting these observations, since the biological mechanisms and clinical implications are not yet clear^22^. In addition, in our analysis, there was no significant association between MASLD and HBV viral load after adjusting for age, ethnicity, nucleus(t)ide analogue (NA) treatment, HBeAg status and sex, which reduces our confidence in the observation.

### Strengths and limitations

A strength of our study is that it adds data to this clinically important field with a well characterised, diverse population. A large proportion of CHB literature comes from Asia, with an under-representation of many other regions with a high burden of CHB, including the WHO African region^31^. Our central London clinic study setting specialises in providing healthcare for individuals from inclusion health populations, particularly vulnerable migrants, who have travelled from many different countries, experience multiple barriers to healthcare, are under-represented in clinical studies, and are disproportionately affected by CHB^32^. The increasing representation of global diversity is important for understanding outcomes in people living with CHB, which may be impacted by variation in HBV genotype, host genetics and environmental factors^31^.

The main limitation of our study is the cross-sectional design, limiting our ability to interpret a causal relationship between MASLD and CHB outcomes. Analysis of the MASLD/CHB interaction is complex, with multiple confounding factors and composite variables, which may mask heterogeneity between individual components (e.g. of the MASLD definition) and therefore limit interpretation of results. Similar to many routinely collected clinical datasets, data missingness is a limitation of this study. Ethnicity data was incomplete which reduced the power of our linear regression models, since it was included as a confounding factor. In addition, some participants did not have complete CMF data, which may have underestimated the number of people with CMFs and MASLD.

### Implications for practice

This study adds valuable data to the growing body of evidence that cardiometabolic comorbidity needs to be considered in the management of CHB. Widening the multi-disciplinary team to include dieticians and peer support workers, ensuring clinicians are confident in CMF assessment and management and providing accessible, culturally relevant patient educational materials on MASLD and CMFs for patients are important goals for holistic CHB care. This concept of making every healthcare contact count is particularly important for people experiencing multiple barriers to accessing healthcare (i.e. inclusion health populations) ^32^.

## Conclusion

In conclusion, MASLD and CMFs are common in people living with CHB, and may influence liver disease outcomes, highlighting the need for integrated approaches to patient care. However, given the complexity of these interactions and potential limitations in causal interpretation, further research is needed to better define these relationships.

## Data Availability

All data produced in the present study are available upon reasonable request to the authors.

## Acknowledgements

We would like to acknowledge the patients involved in this study and all the Mortimer Market Centre viral hepatitis clinic team, including dietitian Damon Nicholls, who are essential to keep the clinic running smoothly, but who have not been directly involved in this study.

## Authors’ contributions

The article idea was conceived by E.M and P.C.M. E.M., P.C.M, A.A-P and A.C. contributed to study design. Data collection was completed by E.M., C.M. and S.O. The manuscript draft was undertaken by E.M. supervised by P.C.M. E.M., C.M., A.K., D.N., S.O, I.G., D.P, E.T., R.G. S.F. A.C., D.M., A.A-P and P.C.M contributed to manuscript review and editing.

